# Infant and Young Child Feeding in the Context of HIV: An Exploration of Barriers in Exclusive Breastfeeding Practice in Dar Es Salaam, Tanzania

**DOI:** 10.1101/2023.12.21.23300402

**Authors:** Goodluck Augustino, Amani Anaeli, Bruno F. Sunguya

## Abstract

**Background:** Ensuring optimal nutrition through early breastfeeding is vital for infant mental development and overall health. HIV infections complicate decisions regarding exclusive breastfeeding, jeopardizing effective infant and young child feeding, which affects nutrition and health outcomes. Recognizing the lack of evidence on barriers to infant feeding in the context of HIV in Tanzania, this study was conducted to explore individual, household, and community obstacles in the Ilala district, Dar es Salaam.

**Methods:** The study used a hospital-based qualitative approach, focusing on HIV-positive mothers with infants aged 3-6 months in Dar es Salaam city. This involved reviewing mothers’ files, conducting interviews with them, and interviewing Reproductive and Child Health clinics (RCH) and community healthcare providers. In total, 27 In-depth interviews were conducted until data saturation was reached, and thematic analysis was used to analyze collected data.

**Findings:** The study identified various barriers to exclusive breastfeeding, encompassing individual factors like work schedules, postpartum depression, and breast conditions. On the household level, barriers included limited access to resources, family influence, and HIV status disclosure reluctance due to stigma. In the community, low retention in the Prevention of Mother-to-Child Transmission (PMTCT) programs plays a pivotal role in hindering exclusive breastfeeding support for HIV-positive mothers.

**Conclusion and Recommendations:** HIV-positive mothers face diverse barriers ranging from individual, household, and community-based barriers. Policies supporting breastfeeding, early detection of postnatal depression and breast problems, and peer support for young mothers are of paramount importance. Food insecurity and HIV stigma should be tackled through income-generating activities, family involvement in PMTCT programs, and awareness campaigns. Community-based counselors play a crucial role in supporting HIV-positive mothers in their exclusive breastfeeding journey to improve PMTCT care retention.

## INTRODUCTION

Optimal nutrition for infants is vital for their development, particularly during the critical first 24 months of life. The period sets the foundation for healthy growth and development, impacting future generations (1). The World Health Organization (WHO) recommends Exclusive Breastfeeding (EBF) for the first six months, complementary feeding for up to 12 months, and continued breastfeeding for up to 24 months (2). However, the estimated rate of EBF is 41% in Sub-Saharan Africa (SSA), with wide regional variations (3). Despite an average breastfeeding duration of 14–19 months in SSA, only 30% - 46% practice EBF for the recommended six months (4–6). In Tanzania, while the EBF rate has improved to 59%, challenges persist, making it crucial to understand the dynamics influencing breastfeeding practices (7).

HIV/AIDS has complicated infant and Young Child Feeding (IYCF) practices. Although the benefit of breastfeeding outweighs the risks even for children exposed to HIV, far fewer children exposed to the infections are exclusively breastfed in fear of new infections. In 2017, an estimated 360,000 children were newly infected with HIV globally, and a significant portion of these infections occurred during breastfeeding (8). While the combination of ART and EBF has proven effective in reducing HIV transmission to less than 1% (9), challenges arise in maintaining consistent ART with EBF breastfeeding (10). Unlike high-income countries where breastfeeding is discouraged postnatally, low and middle-income countries face the dilemma of unsafe formula feeding, leading to higher mortality rates in children under five (11).

In the context of HIV, adherence to the recommended duration of EBF remains a challenge (12). Inconsistent EBF practices among HIV-positive mothers have been observed across SSA countries, reflecting the need for tailored interventions(13–16). Tanzania, despite progress, faces barriers leading to varied EBF rates, such as the decline from 85% at two months to <30% by four months in some areas (17,18). Factors contributing to these challenges include socioeconomic, cultural, and health-related barriers, emphasizing the need for a comprehensive understanding of the contextual influences on breastfeeding practices (12,17–21).

While various studies have explored factors influencing exclusive breastfeeding practices, limited attention has been given to understanding barriers specific to HIV-exposed infants in Tanzania. Existing research has primarily focused on economic aspects, leaving gaps in addressing cultural and societal norms impacting EBF (12,21). This study aims to fill these knowledge gaps by investigating barriers to EBF among HIV-positive mothers according to WHO guidelines in Tanzania. By employing an integrated model of behavioral prediction (22), we aim to provide insights into the determinants affecting exclusive breastfeeding practices, paving the way for targeted interventions.

## METHODS

### Research design and setting

This study adopted a qualitative case study design to explore barriers to exclusive breastfeeding practice among HIV-positive mothers. The selection of a case study design was deliberate, driven by the fact that the chosen diverse cases represented real, ongoing, and current scenarios (23). Many controls on infant feeding choices are knotty and complex to quantify (24). Therefore, using a qualitative method provided a more profound comprehension of the factors that impact infant feeding behaviors and intentions.

The study was conducted in the Ilala district in Dar es Salaam. The region was chosen as it is Tanzania’s largest city, industrial center, and major port (25). In addition to the significant population with mixed cultures from all over and outside the country, the urban-rural migration status in Dar es Salaam is higher than in other regions in Tanzania (25,26). Further, the city is projected to grow beyond 10 million by 2029 and emerge among megacities (26). Urbanization’s influence on breastfeeding and HIV is well established (1,27,28).

The study was done in the highest client flow accessing HIV services through HIV Care and Treatment Centers (CTC) and Reproductive and Child Health (RCH) clinics in Ilala, Amana Regional Referral Hospital, Mnazi Mmoja, and Buguruni Health Centre. Although the regional proportion of EBF is not known among HIV-exposed infants, the overall estimate of EBF in Dar es Salaam among children in the general population is 57.1% (29).

### Recruitment and selection of study participants

Purposive sampling was used to recruit HIV-positive breastfeeding mothers, healthcare providers, and Community Health workers in the Ilala district in Dar es Salaam. HIV-positive mothers who had infants aged 3–6 months, regardless of their current feeding practice, have been placed on a treatment regimen (ARV medicines), attending CTC and RCH between March 08^th^ to 25^th^, 2022, recruitment period of the study were eligible. This age range was chosen for the study based on previous research findings that mothers in Tanzania typically discontinue breastfeeding or begin introducing complementary foods before or around the time their infants reach three months (7,17,30).

Further, a hospital document review was conducted to support a judgmental sampling procedure based on the diversity of sub-categories to identify the participants meeting the sampling criteria. These sub-categories include socio-demographic status (age – Adolescents, above 18 years, educated (at least standard 7 of primary school), uneducated, parity (first child-parent, multiple children), marital status (polygyny, Monogamy, single mother), infant’s gender, and employment status (employed, unemployed) as they have been proven to impact exclusive breastfeeding practice in women in the general population (31). Additionally, clinicians and community health workers from CTC and RCH of the selected health facilities were purposively recruited. Study staff approached eligible healthcare providers and HIV-positive mothers in person at the RCH clinics and asked for an interview after briefing them about the study. A total of 27 interviews was adequate to reach saturation of the data collected in the selected health facilities, including 21 in-depth interviews with HIV-positive mothers, 4 healthcare providers, and 2 community healthcare workers key informant interviews. All eligible participants approached agreed to participate, and no repeat interviews were conducted.

### Data collection and tools

Before collecting data in the selected health facilities, the interview guides underwent a rigorous pilot testing phase outside the selected study sites at Tabata Health Center. In-depth and key informant interviews were meticulously conducted within a private room adjacent to the RCH building, involving only the interviewee and the participants. Following each interview, comprehensive field notes were diligently recorded. The duration of interviews varied between 20 to 40 minutes per session. Notably, data saturation was observed after the 18th interview, as much of the information gathered exhibited a repetitive pattern.

### In-depth Interviews

A document review was conducted to elicit background information for the interview. It was done purposely to collect the participants’ socio-demographic data, which was crucial to obtaining the desired number of study participants. Whereas the regional and district authorities sought permission to acquire the documents, confidentiality at the individual level was ensured before the interviews. Required records were compiled, and a brief understanding of their production was provided. Accuracy was determined by comparing the social and demographic data among the documents, and the summary was created to assist in sample selection for in-depth interviews. Employing comprehensive interviews to investigate how infants are fed in the context of HIV is consistent with the methodology used by researchers who conducted comparable studies in SSA (32). In-depth interviews elicited in-depth responses, which enabled the interviewer to fully explore the reasons, feelings, beliefs, and opinions that highlight individual participants’ responses (33) since infant feeding is personal and rooted in the cultural and social norms of the community (34).

### Key Informant Interviews

This was used to generate information from healthcare providers at the selected clinics in the health facilities. In addition, we included community health workers from respective healthcare facilities. These are thought to be information-rich regarding exclusive breastfeeding practices among HIV-positive women.

### Data Management and analysis strategies

Managing the data began in the field by ensuring that all participant observations and reports were recorded correctly and appropriately. This involved ensuring that codes were assigned accurately and validly. The study utilized the thematic analysis technique to derive meaning from the data collected and analyze it based on the research objective and questions.

After collecting the data, the researcher read and reread the entire dataset to understand its depth and breadth. The Nvivo software was used during the analysis, and five steps of qualitative data analysis framework were adopted as follows: i. Verbatim transcribed ii. Familiarize myself with data by reading each transcript and writing labels to the data. iii. Identifying a thematic framework by jotting down key concepts re-emerging from the data iv. Deductively indexing the data by grouping them according to the open codes presented in conceptual framework v. mapping and interpretation of the data through which data were examined and interpreted (23)

Facts and participants’ views were presented in “quotes,” which were identified by the participant number and health facility name, and the third-person voice was used to analyze the data while reporting the data collected. No personalized ideas were included in the final report, and the facts were presented through the actual words of the participants. The objective data from in-depth interviews were sequentially coded and reported objectively. Furthermore, consultation with other social science researchers was sought to gain different perspectives on proper data management and analysis.

### Ethical consideration

This study received ethical approval from the Research Ethics Committee of Muhimbili University of Health and Allied Sciences (Ref. No. DA.282/298/01.C/MUHAS-REC-08-2021-813). Additionally, research clearance was obtained from the Dar es Salaam Regional Authority and Ilala Council Authority. Written consent was diligently obtained from all participants, accompanied by a clear assurance of their right to withdraw from the study at any point. Maintaining confidentiality and respecting privacy were paramount, with no names being recorded in the transcripts.

## RESULTS

The findings of this study are structured to reflect the research questions and objectives on the central theme of barriers to exclusive breastfeeding among HIV-positive women attending clinics in the study area. The results were initially grouped into individual, household, and community themes. The themes are formed with sub-themes illustrated in Table 1 below. A verbatim quotation from the collected data supports each sub-theme. The verbatim inclusion in presenting research findings is crucial to demonstrating evidence and the data generation process (35). In addition, the validity and reliability of the conclusions presented through “detailed and thick verbatim descriptions of participants’ accounts to support findings.” Themes and sub-themes related to the barriers to EBF are summarized in the table below and are detailed in subsequent paragraphs.

**Table 1:** Summary of Theme and Sub-themes.

| Theme number | Themes | Sub-themes |
| --- | --- | --- |
| Theme 1 | Individual barriers to exclusive breastfeeding | Occupation-related hectic schedule |
|  |  | Early motherhood-related non-compliance to the safe infant feeding directives |
|  |  | Postpartum depression |
|  |  | Breast sores and abscess |
| Theme 2 | Household-level barriers to exclusive breastfeeding | Food insecurity and inaccessibility to key resources |
|  |  | Male partners and family members influence on decision making |
|  |  | HIV status non-disclosure barriers to EBF support |
| Theme 3 | Community-level barriers to exclusive breastfeeding | Low retentivity of HIV-positive women in PMTCT programs |

### Theme 1: Individual barriers to exclusive breastfeeding

Participants in this study described individual barriers faced during exclusive breastfeeding. Under this theme, four sub-themes emerged, including early motherhood-related non-compliance to the safe infant feeding directives, occupation-related schedule, postpartum depression, and breast sores and abscesses. The sub-themes are detailed below with support of verbatim quotations.

### Occupation-related schedule

Our study reveals that strict rules of mothers’ occupation have significantly influenced mixed feeding. The work that capacitated them to provide for themselves from the income generated made mothers trade off caring for their infants over their jobs. As a result, they neither offer maternity leave, mid-day time out to breastfeed nor reduced work hours to breastfeed their infants, and self-caring to produce enough milk for their infants. One of the participants, who shared their valuable insights, highlighted a challenging situation. This participant said:

> *“I had to stop my child from breastfeeding and leave him with soft porridge since I am working as a maid, where we work 12-hour shifts.” (P12, Health Center)*

This statement underscores the impact of employment on exclusive breastfeeding practices. The participant’s work schedule and responsibilities made it logistically challenging to breastfeed their child during mid-day. Furthermore, the participant expressed concerns about potential risks associated with the child’s caretaker. She explained the following:

> *“Moreover, I was afraid that when I was not around, my young sister, whom I left the baby with, might be tempted to give him sweet drinks like Soda to calm him; when my child gets irritated and crying, that might be more dangerous.” (P12, Health Center)*

This apprehension indicates the awareness of the importance of exclusive breastfeeding and the potential consequences of introducing non-breast milk items to the infant’s diet.

We also gained valuable insights from a community health worker (CHW) behind mixed feeding practices among mothers, which are often influenced by the mothers’ occupations, particularly those working in industry-related jobs and Hindu facilities. The main challenge for these women is the need for more time off to breastfeed their infants frequently. The participant said:

> *The most reason we encounter when trying to probe mixed feeding practice is influenced by their occupation, which comes from these industry-related occupations and those working in Hindu facilities not being given time off to breastfeed their kids. Moreover, our clients have yet to seriously consider the advice of expressing breastmilk and leaving it at home. (PRC1, Health Center)*

Further, there was limited understanding of how to extract breast milk, which was perceived as unnatural unless the mother was hospitalized. This uncommon practice in the community resulted in a constraint on exclusive breastfeeding. There were also apprehensions about expressing and storing milk, sanitation, and the notion that it conflicted with God’s established values. The participant said:

> *I have tried to express breast milk several times, but it has never been suitable for my child. It feels like he gets more abdominal discomfort when I give them to him (P21, Health Center)*

### Early motherhood-related non-compliance to the safe infant feeding directive

It was reported that most adolescent-age mothers were reluctant to follow the advice provided during the clinics and visits from CHWs. It was termed as ignoring the child’s health outcomes, still feeling irresponsible for their child’s health, and further deciding to adhere to their parents’ and community’s inexpert opinions on alternatives to breastfeeding. Stressing on this matter, the participant said:

> *After realizing that I had a pregnancy and was diagnosed with HIV at 19, the home was no longer safe. So, I moved to live with my best friend (co-worker), who has been by my side for better or worse. I trust and share everything with her, even her opinion on feeding my child since she has two children and is more experienced (P22, RRH)*.

The insight from a CHW revealed that knowledge uptake on infant and young child feeding was generally high among parents aged 24 and older. However, the CHW identified a challenge in getting younger parents to translate knowledge into practice, particularly in the 16-24 age group. While they may have verbally expressed willingness to follow health teachings, their child’s growth did not consistently reflect this commitment. Instead, these younger parents often relied on peer opinions for guidance. The participant said:

> *Knowledge uptake, in general, is high, but only above 24 years of age, mainly under that age, it has been challenging to uptake knowledge that we give to them. They will lie to follow our teachings, but it does not reflect on the child’s progress, mostly being careless of their child’s growth; instead, they rely on peer opinions. The leading age group we struggle with is 16-24 years (PRC12, Health Center)*.

This observation underscores the need for tailored approaches and interventions to effectively engage and educate younger parents on optimal infant and young child feeding practices.

### Postpartum depression

Postpartum stress/depression fueled by the partner and mother’s family emerged among the barriers that hinder mothers from practicing EBF, mainly reported an unhealthy relationship with their partners. Further, there is a lack of support for the different phases of motherhood from the partner aggravated with newly diagnosed HIV during the ante-natal clinic. As a result, giving up and neglecting self-health measures, including food intake, leads to a massive reduction in milk production and disinterest in breastfeeding the child. Describing this situation, a participant said:

> *I would be lying to say I hate this child, but I would instead focus on my small business since no one has my back to provide for my other older child and me. I have been suffering a lot from pregnancy with no support, and now I must raise this child alone. I know the effect of alternative feeds, but I don’t believe it is my fault alone. If I cannot produce enough milk, there is no way I will leave him hungry with no food (P16, Health Center)*.

### Breast sores and abscess

The health care provider reported sporadic cases, and for a few days (Five), as notified by one of the mothers interviewed, presented with pustules during the early days of breastfeeding, mostly treated and recovered within a short time with no recurrences. The participant said:

> *In the second month of breastfeeding, my left breast developed something like an abscess. I immediately went to the clinic and was treated with medication and told to breastfeed on one side only and rest the affected breast while expressing the breast milk from it and dumping them. I recovered shortly without complications (P17, Health Center)*.

### Theme 2: Household-level barriers to exclusive breastfeeding

Participants also reported household-level barriers faced during exclusive breastfeeding practice. In this theme, three sub-themes, including food insecurity and inaccessibility to key resources, male partners and family members’ influence on decision-making, and HIV status non-disclosure barriers to EBF support, are presented below. Verbatim quotations support the presentation.

### Food insecurity and inaccessibility to key resources

Participants in this study revealed that their limited access to food was a primary reason for their inability to breastfeed their children exclusively. Due to insufficient meals (typically one to two per day), mothers produced inadequate breastmilk, which failed to satisfy their infants, and sometimes, they had no breastmilk at all. This food insecurity led them to seek help from relatives when possible, but maintaining consistent access to food for themselves and their infants remained challenging. Occasionally, this was reported, leading to considerations of alternative feeding practices.

> *The main reason I failed to EBF my child was food, with one to a maximum of two meals per day leading me to produce very light breastmilk, which does not satisfy my child, and sometimes no breastmilk at all. So, I sometimes must go to my relatives to ask for food for its sake. However, I am not consistently successful; when I have no alternative for feeding myself, I consider something else for my child (P31, Health Center)*.

Further, participants shared that the demanding nature of breastfeeding, coupled with the need to eat frequently to sustain the energy necessary for both breastfeeding and adhering to ART, constantly left them feeling weak. Despite their consistent efforts, the situation did not improve. Consequently, the participant made the difficult decision to cease breastfeeding altogether and transition to providing complete replacement feeds such as lactogen to their child.

> *Breastfeeding was hard for me, and I was always weak. This is because I had to eat many times daily to maintain the energy required to breastfeed and ART intake but the situation remains the same. Hence, I decided to stop for good and provide her with complete replacement feeds through lactogen, which significantly relieved my health and hers (P14, Health Center)*.

These accounts highlight the intricate balance required for mothers living with HIV who are striving to breastfeed while managing their health through ART and the difficult decisions they often face.

A participant from a healthcare facility reported that a common concern among clients is poor nutrition, which affects a substantial portion of them. This nutritional deficiency often results in insufficient breast milk production, compelling many of these mothers to seek more affordable alternatives, with soft porridge being a common choice.

> *Most of our clients face poor nutrition; I would say 6 out of 10 clients. That leads to insufficient breastmilk production and the choice of soft porridge as their most affordable alternative (PR2, RRH)*.

### Male partners and family members have bold authority in decision-making

In this sub-theme, most of the participants in the study reported taking a leading role in feeding their infants. While only a few said their partners fulfilled that rule. Our participants presented the dominance of family members in making decisions on infants while ignoring or not seeking advice from the experts in health facilities. One of the participants said:

> *I was not advised to feed my child from the clinic. I went there during pregnancy and after delivery; hence, I relied on my husband’s advice. We fed him cow milk with soft cassava porridge when he was three months old*. *My child cannot feed on only breastmilk. Even those doctors’ infants cannot sustain breastmilk alone, even without water! No way (P32, Health Center)*

Couples who were not willing to undergo HIV testing or visit the clinic showed a decreased tendency to value women’s choices of exclusively breastfeeding their infants. In addition, most male partners seemed to lack vital information on their infants’ proper feeding, hence improper influence on their wives.

### HIV-associated stigma, disclosure about their HIV status

The findings revealed a high level of undisclosed HIV status among the co-parents and other close persons. A ratio of 4:10 of the participants had not disclosed their status to their partner. This negatively affects the practice of EBF despite the knowledge disseminated by the health care providers. HIV-positive mothers reported positive support from their spouses in ensuring food security for sufficient milk production and replacement food for those who opted to use complete replacement feeding (CRF) when becoming aware of their HIV status.

> *My husband has been enormous support from the day I was diagnosed with HIV, during the clinic of this child. I lost hope and even wanted to abort. However, despite being tested negative, he did increase his love and affection for me, and to date, he has been of hope and colossal support that I need in every way, from my health, feeding pattern, and protection for the child’s health. We attend all the counseling sessions together and support each other to protect our child’s health (P19, Health Center)*

Participants reported supporting their family members and partners’ mixed feeding opinions to continue concealing their HIV status despite the special consideration required by their exposed children.

A notable trend has emerged in our research, as reported by CHW. Male partners who are aware of their wives’ HIV status have demonstrated a commendable level of involvement. This heightened male-partner engagement is positively reflected in the practice of exclusive breastfeeding (EBF) among these couples. Additionally, it has translated into active participation in community outreach programs, underlining the importance of involving male partners in promoting EBF and broader community health initiatives.

> *We have had an outstanding response from male partners who know their wives’ status. It has been reflected in the EBF practice among those couples and significant participation in community outreaches (PRC12, Health Center)*.

### Theme 3: Community-level barrier to exclusive breastfeeding

Participants reported community-level barriers faced during exclusive breastfeeding practice. In this theme, only one sub-theme emerged, which is the Low retentivity of HIV-positive women in PMTCT programs. This is substantiated below with the support of a verbatim quotation.

### Low retentivity of HIV-positive women in PMTCT programs

As reported by participants, keeping women and infants in PMTCT programs after delivery has been challenging. In addition, there has been gradual progress in HIV-positive mothers’ detachment from postnatal clinics and community outreaches targeting PMTCT services. With a range of benefits to women and infants, one of the key supports of PMTCT services is safe childbirth and appropriate infant feeding. One of the healthcare providers said:

> *As soon as they finish the cluster of vaccines given during the first three months, the number of HIV-positive mothers attending PMTCT clinics and community outreaches significantly reduces. You can imagine what comes next: reducing infant care and improper feeding. Unfortunately, most dropouts are those coming from the neighborhood, and they do so to conceal their status (PR1, Health Center)*

As a result, difficulty in reducing, detecting, and managing new infections among HIV-positive mothers throughout breastfeeding predisposes them to drop EBF practice. This observation underscores the need for tailored interventions and supports to retain these mothers in PMTCT services and ensure the well-being of both mothers and infants.

## DISCUSSION

This study explored the challenges HIV-positive mothers face in practicing exclusive breastfeeding (EBF) at different levels. It revealed individual, household, and community barriers that hinder the practice of exclusive breastfeeding (EBF) among HIV-positive mothers. Individual factors, such as occupation tie, early motherhood, postpartum depression, and breast conditions like sores and abscesses, were reported. Household challenges encompassed issues such as food insecurity, the impact of spouses and family authority on decision-making, and a lack of substantial support from significant others due to HIV-related stigma. Notably, disclosure hesitance and HIV-related stigma emerged as intertwined factors in this complex scenario. Additionally, key informants highlighted a community-level barrier: the low retention of HIV-positive mothers in PMTCT programs.

The voice of our participants in quotes from the results section sheds light on the critical issue of occupation as a significant barrier to exclusive breastfeeding (EBF) among HIV-positive mothers. HIV-positive mothers are often required to balance their occupational responsibilities with childcare, thus encountering substantial hurdles in ensuring EBF. The demands of the workplace can frequently disrupt breastfeeding schedules, leading to suboptimal feeding practices. This observation aligns with the broader literature on working mothers’ challenges sustaining EBF. Employment can positively impact breastfeeding as it provides financial stability and a sense of purpose, allowing mothers to invest in their health and the health of their infants (36).

On the contrary, employment can also negatively affect breastfeeding, as mothers reported being unable to breastfeed exclusively due to work schedules, lack of appropriate facilities, or stigma surrounding HIV and breastfeeding, similar to other contexts (37). Employed women are more likely to practice EBF than unemployed women (38). Some of the reasons, such as employment-related stress and lack of flexible working hours, are reported to be associated with lower rates of EBF among HIV-positive mothers (39). Providing workplace support, including designated spaces for breastfeeding, lactation breaks, and education on the benefits of exclusive breastfeeding, has the potential to increase exclusive breastfeeding rates among working mothers and can also positively impact the exclusive breastfeeding rates among HIV-positive mothers. (40,41)

The age of mothers plays a crucial role in their ability to adhere to exclusive breastfeeding (EBF) practices, particularly among HIV-positive mothers. Despite high knowledge uptake on EBF as reported by healthcare providers, a notable distinction arises when considering age groups. Mothers aged 24 and above tend to have a more significant understanding and commitment to EBF, while those under this age, especially teenagers, face challenges in adopting the knowledge provided. Adolescent-aged mothers were pessimistic towards EBF due to the peer influence from their age mates and parents, making it difficult for them to initiate and sustain exclusive breastfeeding, similar to other contexts (11). Mothers often report being advised by their families to use mixed feeding for their infants, even if they disagree, primarily if they rely on their families for support (42). Adolescent mothers who are HIV positive might lack experience and confidence in their beliefs, which could lead them to seek assistance with parenting from their family members, especially their mothers and grandmothers (37). This is particularly true for adolescent mothers who are HIV-positive and may feel unsure about their parenting abilities (37). They may turn to their mothers and grandmothers for guidance, and accommodating their families’ wishes may help them cope with the demands of parenting while still developing themselves (42). However, there is inconsistency in the findings of different studies. For example, younger mothers had higher exclusive breastfeeding rates than older mothers, as reported elsewhere, and HIV-positive mothers under 25 had higher exclusive breastfeeding rates than those over 25 (38,43). It is, therefore, essential to note that a mother’s age is not the sole determinant of exclusive breastfeeding practices (44). Other factors, including access to healthcare services, education, and social support, can also influence exclusive breastfeeding rates (6,44).

This study demonstrates that mothers who experience emotional breakdowns and stress are at a higher risk for poor self-efficacy and may neglect their infants’ care. Unhealthy family relationships, loneliness, and feelings of rejection can lead to severe stress and depression. Thus, mothers who lack psychosocial support may struggle with the challenges associated with exclusive breastfeeding (EBF) and experience guilt and feelings of inadequacy. Numerous studies have indicated that social and family support quality can positively impact neuroendocrine functioning and mood (45–49). These emotions are often linked to early discontinuation of breastfeeding and exacerbate the connection between maternal stress and psychological distress (50,51). Prenatal depression also significantly affects EBF rates, with studies reporting poor rates in South Africa and Australia (52). Therefore, the presence of depression during pregnancy in women with HIV poses a risk to their capacity to effectively follow the World Health Organization’s guidelines, which include exclusive breastfeeding for up to 6 months and adherence to antiretroviral therapy. A significant number of self-reported depression within six months post-delivery reported in Australia pose more threat to EBF discontinuation and hence require more attention (44). Women with more depressive symptoms were less likely to initiate breastfeeding exclusively, as noted in previous studies of depressive symptoms and poor infant nutrition (53). Further, women with depressive symptoms are less likely to start and maintain exclusive breastfeeding, as seen in studies from Kenya and other countries. (52,54)

Breast conditions, such as abscesses, mastitis, or other infections, pose a considerable threat. In response, healthcare providers often recommend limiting breastfeeding from the affected breast while expressing and discarding the milk to prevent potential transmission of the virus. While such interventions can help mitigate risks, they introduce additional burdens and complexities for mothers. HIV-positive mothers are advised to stop breastfeeding and seek immediate treatment while using alternative feeds that are safe for their infants’ such as safe formula feed (55,56). However, the cost and the competitive price make it difficult for them to sustain for a slightly extended period, hence including cow milk among the options (57). HIV infections create a significant risk of developing breast infection (58), and the risk is reduced with the effective use and adherence to potent ART. The same findings were reported in Ghana, where some participants identified improper positioning of the baby as the cause of sore nipples (59). In addition, incorrect techniques, less frequent breastfeeding, scheduled times, and pacifiers were reported to predispose to lactation problems (60). Breast-related conditions are preventable if the mother empties her breasts and effectively starts post-delivery immediately and beyond. Properly managing these conditions is crucial; if not treated, it might lead to early weaning.

The study findings illustrate that mothers facing food insecurity often struggle to provide sufficient and nutritious breastmilk to satisfy their infants. The struggle to nourish their infants while grappling with their hunger adds a layer of stress to their lives. This emotional burden can negatively impact the mother’s well-being, potentially affecting her capacity to engage in EBF. Inadequate nutrition can have a notable impact on the physiology of the breast and the production, secretion, and composition of milk (61). In addition, the infant’s fussiness and poor weight gain force them to supplement their breast milk with other food sources and, as a result, reduce the benefits of exclusive breastfeeding for both the mother and the baby. Similar associations between food insecurity and negative attitudes toward EBF have been found in other contexts (39,40). In Malawi, food insecurity was a significant factor in HIV-affected women’s decisions to continue breastfeeding (10). Women often believe inadequate diets lead to insufficient milk production, increasing the likelihood of supplementing their infants (13,37,62). Their hunger drives the perception of their bodies inability to produce sufficient milk, leading to a reinforcement of mixed feeding to meet their perceived needs for their children (63).

The social dynamics surrounding infant feeding have a considerable impact on a woman’s ability to maintain optimal breastfeeding practices, regardless of her HIV status. While healthcare providers offer guidance to address household obstacles, such as pre-lacteal feeds or herbal remedies and water, mothers-in-law and husbands, who play an essential role in supporting mothers in the domestic sphere, often provide erroneous advice regarding infant feeding. Consequently, their understanding of infant feeding becomes the primary source of information for women in this community. This study is in line with the systematic review of the views, attitudes, and behaviors of both individuals and societies concerning mother-to-child transmission and infant feeding among HIV-positive mothers in SSA (64), demonstrating that household culture and social norms influence a mother’s decision to EBF (64). In addition, interventions encouraging male partner involvement, such as enhanced psychosocial interventions, verbal encouragement, and complex community interventions, increase safe infant feeding practices.

The act of revealing one’s HIV status is a complicated matter since it carries crucial psychological, social, cultural, and financial consequences. In this study, participants who reported not disclosing their HIV status to anyone, including their partners, were most likely to favor mixed feeding practices. In contrast, women who declared their status reported massive support from their close people, which influences EBF practice and collective support for infant feeding. Healthcare providers also commented on the positive impact of the disclosure among the couple that disclosed to each other on adherence to the infant feeding recommendations and enabling her to take medication without fear. This finding agreed with the study conducted in Nigeria (65). Further, EBF practices among HIV-positive mothers who disclosed their status to their spouses were nearly six times more likely than mothers who did not disclose their status in Ethiopia (66). The possible reason would be that if mothers declare their HIV status to their spouses, they might get good family care. For instance, some reasons for disclosure were decreased workload, nutritional reinforcement, compliance with ART treatments, and exclusive breastfeeding motivation (13,67). Moreover, disclosing HIV status to the spouse prevents mixed feeding (66). This is because HIV-positive mothers who tell their status to their partner will get adequate care, support, and time for breastfeeding.

Poor retention of HIV-positive mothers to PMTCT services, which starts from the Antenatal clinic (ANC), was also reported to be associated with reduced EBF practice in our study despite the local availability of health facilities. The main reason for poor clinic attendance was women’s desperation to conceal their HIV status from close family members. This compels them to stay away from the facilities where HIV status was diagnosed, where they can receive PMTCT services, and attend perinatal clinics where status is unknown. Several studies have reported the positive contribution of PMTCT services to EBF practice among HIV-positive mothers (68–70). As proof, PMTCT services improved knowledge, attitudes, and exclusive breastfeeding practices among HIV-positive mothers (68–70). Further, PMTCT services help educate and support mothers to practice exclusive breastfeeding which is crucial in reducing the risk of mother-to-child virus transmission and providing essential nutrients to the infant (69). The challenge for women to disclose their serostatus and the stigma associated with it creates obstacles to their engagement in PMTCT programs and starting or continuing ARV therapy for their well-being and that of their children (71,72). Consequently, they are also less inclined to adopt exclusive breastfeeding practices (71,72). However, women are concerned about the implication of close relatives becoming aware of their positive status. Augmenting essential PMTCT services with mentor mothers and a culturally adapted cognitive behavioral intervention helped give information and improve the emotional perspective of HIV-positive pregnant women (73). Hence, healthcare providers should mount efforts to enhance the effectiveness of PMTCT programs and identify and involve these significant others to help disperse the stigmatization of HIV-positive women.

### Study limitations and strengths

We ensured transparency in the study’s objectives before interviews to mitigate the potential impact of social desirability bias, especially in discussions about infant feeding practices. Participants were explicitly informed of the study’s purpose and reassured that there would be no adverse consequences related to their chosen feeding methods. Furthermore, using Kiswahili, the national language, during data collection facilitated open and candid communication, allowing participants to express themselves comfortably in their native language, which aided in code identification. Additionally, to enhance the trustworthiness of our qualitative research, we employed data triangulation involving three distinct participant groups and utilized two researchers for transcript coding. Including direct quotes from participants describing their experiences further enhances the reliability of the study findings, providing readers with concrete evidence of the study’s dependability.

## CONCLUSIONS AND RECOMMENDATIONS

This study sheds light on the intricacies of optimal breastfeeding, specifically exclusive breastfeeding as a method of infant feeding for HIV-positive mothers. The results indicated several barriers to exclusive breastfeeding (EBF) among these mothers. Individual factors, such as a busy work schedule, early motherhood, postpartum depression, and breast conditions such as sores and abscesses, were significant challenges to EBF. Household challenges, including food insecurity, the influence of spouses and family authority on decision-making, and lack of substantial other support due to HIV-related stigma and disclosure hesitance, hindered EBF practice. HIV-related stigma and disclosure hesitance were also barriers to EBF. Furthermore, low retention of HIV-positive mothers in PMTCT programs emerged as a significant obstacle to EBF practice in the community. These findings suggest a high need for a more individualized, household, and communalized approach to sustaining HIV-positive mothers to adhere to exclusive breastfeeding.

Authors’ contribution: GAM conceptualized the research questions, developed the proposal, conducted data collection, analysed data, and prepared the first draft manuscript; AAM developed the proposal, and tools, supported analyses, and edited the manuscript; BFS supervised the proposal development, supported analyses, and edited the manuscript.

## Data Availability

If the data are all contained within the manuscript and/or Supporting Information files, enter the following: All relevant data are within the manuscript and its Supporting Information files.

## REFERENCES

1. UNICEF. United Nations Children’s Fund. Progress-for-every-child-in-the-sdg-era. New York. New York: UNICEF; 2019 p. 1–13.

2. World Health Organization. Updates on HIV and infant feeding Guideline. Geneva: WHO; 2016 p. 1–68.

3. World Health Organization. Enabling women to breastfeed through better policies and programs: Global breastfeeding scorecard. WHO; 2018 p. 1–4.

4. Bbaale E. Determinants of early initiation, exclusiveness, and duration of breastfeeding in Uganda. J Health Popul Nutr. 2014 Jun;32(2):249–60.

5. Traoré M, Sangho H, Camara Diagne M, Faye A, Sidibé A, Koné K, et al. Facteurs associés à l’allaitement maternel exclusif chez les mères d’enfants de 24 mois à Bamako. Santé Publique. 2014;26(2).

6. Kimani-Murage EW, Wekesah F, Wanjohi M, Kyobutungi C, Ezeh AC, Musoke RN, et al. Factors affecting actualisation of the <scp>WHO</scp> breastfeeding recommendations in urban poor settings in <scp>K</scp> enya. Matern Child Nutr. 2015 Jul 17;11(3).

7. Ministry of Health Gender Elderly and Children - MoHCDGEC/Tanzania Mainland CD, MoH/Zanzibar M of H, NBS/Tanzania NB of S, OCGS/Zanzibar O of CGS, F IC. Tanzania Demographic and Health Survey and Malaria Indicator Survey 2015-2016. Dar es Salaam, Tanzania: MoHCDGEC, MoH, NBS, OCGS, and ICF; 2016.

8. UNAIDS. United Nations Programme on HIV/AIDS. UNAIDS DATA 2019. UNAIDS; 2019 p. 1–471.

9. Coovadia H, Kindra G. Breastfeeding to prevent HIV transmission in infants: balancing pros and cons. Curr Opin Infect Dis. 2008;21(1):11–5.

10. Kafulafula UK, Hutchinson MK, Gennaro S, Guttmacher S. Maternal and health care workers’ perceptions of the effects of exclusive breastfeeding by HIV positive mothers on maternal and infant health in Blantyre, Malawi. BMC Pregnancy Childbirth. 2014 Dec 25;14(1).

11. Lawani L, Onyebuchi A, Iyoke C, Onoh R, NKWO P. The challenges of adherence to infant feeding choices in prevention of mother-to-child transmission of HIV infections in South East Nigeria. Patient Prefer Adherence. 2014 Mar;

12. Husain Rasheed M, Philemon R, Damas Kinabo G, Maxym M, Mamuu Shayo A, Theophil Mmbaga B. Adherence to Exclusive Breastfeeding and Associated Factors in Mothers of HIV-Exposed Infants Receiving Care at Kilimanjaro Christian Medical Centre, Tanzania. East Afr Health Res J. 2018 Apr 1;2(1).

13. Belay GM, Wubneh CA. Infant Feeding Practices of HIV Positive Mothers and Its Association with Counseling and HIV Disclosure Status in Ethiopia: A Systematic Review and Meta-Analysis. AIDS Res Treat. 2019 Aug 1;2019.

14. Andare N, Ochola S, Chege P. Determinants of infant feeding practices among mothers living with HIV attending prevention of mother to child transmission Clinic at Kiambu Level 4 hospital, Kenya: a cross-sectional study. Nutr J. 2019 Dec 2;18(1).

15. Aishat U, David D, Olufunmilayo F. Exclusive breastfeeding and HIV/AIDS: à crossectional survey of mothers attending prevention of mother-to-child transmission of HIV clinics in southwestern Nigeria. Pan Afr Med J. 2015;21.

16. Napyo A, Tumwine JK, Mukunya D, Waako P, Tylleskär T, Ndeezi G. Exclusive breastfeeding among HIV exposed infants from birth to 14 weeks of life in Lira, Northern Uganda: a prospective cohort study. Glob Health Action. 2020 Dec 31;13(1).

17. Manji KP, Duggan C, Liu E, Bosch R, Kisenge R, Aboud S, et al. Exclusive Breast-feeding Protects against Mother-to-Child Transmission of HIV-1 through 12 Months of Age in Tanzania. J Trop Pediatr. 2016 Aug;62(4).

18. Young SL, Israel-Ballard KA, Dantzer EA, Ngonyani MM, Nyambo MT, Ash DM, et al. Infant feeding practices among HIV-positive women in Dar es Salaam, Tanzania, indicate a need for more intensive infant feeding counselling. Public Health Nutr. 2010 Dec 29;13(12).

19. Saka FJ. “Factors Influencing Exclusive Breastfeeding Among HIV Positive Mothers at Ilala Municipality Dar es Salaam.” MUHAS 2012 Oaiihieprintsorg1662. 2012;

20. Mnongya L. Dilemma of choice between breastfeeding and replacement feeding among HIV positive mothers in Tanzania. Dar Es Salaam Med Stud J. 2013 May 22;19(2).

21. Williams AM, Chantry C, Geubbels EL, Ramaiya AK, Shemdoe AI, Tancredi DJ, et al. Breastfeeding and Complementary Feeding Practices among HIV-Exposed Infants in Coastal Tanzania. J Hum Lact. 2016 Feb 30;32(1).

22. Fishbein M (2009). An integrative model for behavioral prediction and its application to health promotion. Emerg Theor Health Promot Pract Res. 2009;215–34.

23. Creswell JW. Educational research: Planning, conducting, and evaluating quantitative and qualitative research. Vol. 4. Boston, MA: Pearson; 2012.

24. Zulliger R, Abrams EJ, Myer L. Diversity of influences on infant feeding strategies in women living with HIV in Cape Town, South Africa: a mixed methods study. Trop Med Int Health. 2013 Dec;18(12).

25. United Nations Human Settlements Programme. Regional and Technical Cooperation Division. Tanzania: Dar Es Salaam city profile. UN-Habitat; 2009. 32 p.

26. Nations U, of Economic D, Affairs S, Division P. World Urbanization Prospects The 2018 Revision. 2018.

27. Dyson T. “HIV/AIDS and Urbanization.” Population and Development Review. http://www.jstor.org/stable/3115281 [Internet]. 2003 [cited 2022 Jun 22];29(Population Council). Available from: http://www.jstor.org/stable/3115281

28. Yin XH, Zhao C, Yang YM, Shi HF, Wu TC, Xie JL, et al. What is the impact of rural-to-urban migration on exclusive breastfeeding: a population-based cross-sectional study. Int Breastfeed J. 2020 Dec 14;15(1):86.

29. Ministry of Health CD Gender, Elderly and Children (MoHCDGEC) [Tanzania Mainland], Ministry of Health (MoH) [Zanzibar], Tanzania Food and Nutrition Centre (TFNC), National Bu-reau of Statistics (NBS), Office of the Chief Government Statistician (OCGS) [Zanzibar]., UNICEF. Tanzania National Nutrition Survey using SMART Methodology (TNNS) 2018. Dar es Salaam: MoHCDGEC, MoH, TFNC, NBS, OCGS, and UNICEF; 2018.

30. Coovadia HM, Rollins NC, Bland RM, Little K, Coutsoudis A, Bennish ML, et al. Mother-to-child transmission of HIV-1 infection during exclusive breastfeeding in the first 6 months of life: an intervention cohort study. The Lancet. 2007 Mar;369(9567).

31. Maonga AR, Mahande MJ, Damian DJ, Msuya SE. Factors Affecting Exclusive Breastfeeding among Women in Muheza District Tanga Northeastern Tanzania: A Mixed Method Community Based Study. Matern Child Health J. 2016 Jan 4;20(1).

32. Kakute PN, Ngum J, Mitchell P, Kroll KA, Forgwei GW, Ngwang LK, et al. Cultural Barriers to Exclusive Breastfeeding by Mothers in a Rural Area of Cameroon, Africa. J Midwifery Womens Health. 2005 Jul 8;50(4).

33. Barnes BR. Using Mixed Methods in South African Psychological Research. South Afr J Psychol. 2012 Dec 1;42(4).

34. Ijumba P, Doherty T, Jackson D, Tomlinson M, Sanders D, Persson LA. Free formula milk in the prevention of mother-to-child transmission programme: voices of a peri-urban community in South Africa on policy change. Health Policy Plan. 2013 Oct 1;28(7).

35. Patton MQ. Qualitative research & evaluation methods. Sage. 2002;3rd.

36. Frank E. Breastfeeding and maternal employment: two rights don’t make a wrong. The Lancet. 1998 Oct;352(9134):1083–4.

37. Thairu LN, Pelto GH, Rollins NC, Bland RM, Ntshangase N. Sociocultural influences on infant feeding decisions among HIV-infected women in rural Kwa-Zulu Natal, South Africa. Matern Child Nutr. 2005 Jan;1(1).

38. Li R, Liu L, Odent M. Age of mother and exclusive breastfeeding: a systematic review and meta-analysis. Pediatrics. 2016 Apr;137(e20154209).

39. Mundagowa PT, Chadambuka EM, Chimberengwa PT, Mukora-Mutseyekwa F. Determinants of exclusive breastfeeding among mothers of infants aged 6 to 12 months in Gwanda District, Zimbabwe. Int Breastfeed J. 2019 Dec 9;14(1):30.

40. Okereke EI, Ajayi IO, Agho KE, Dibley MJ. Workplace support for exclusive breastfeeding among human immunodeficiency virus-positive mothers. Int Breastfeed J. 2018;13 (1)-15.

41. Vilar-Compte M, Hernández-Cordero S, Ancira-Moreno M, Burrola-Méndez S, Ferre-Eguiluz I, Omaña I, et al. Breastfeeding at the workplace: a systematic review of interventions to improve workplace environments to facilitate breastfeeding among working women. Int J Equity Health. 2021 Dec 29;20(1):110.

42. Bentley M, Gavin L, Black MM, Teti L. Infant feeding practices of low-income, African-American, adolescent mothers: an ecological, multigenerational perspective. Soc Sci Med. 1999 Oct;49(8).

43. Adeyemo TA, Lawuyi OO, Fasubaa OB. Exclusive breastfeeding practices among mothers attending ante-natal clinics in a rural community in Nigeria. J Matern Fetal Neonatal Med. 2018 Apr 31;7(808–14).

44. Forster DA, McLachlan HL, Lumley J. Factors associated with breastfeeding at six months postpartum in a group of Australian women. Int Breastfeed J. 2006;1(1).

45. Brandão T, Brites R, Hipólito J, Pires M, Nunes O. Dyadic coping, marital adjustment and quality of life in couples during pregnancy: an actor–partner approach. J Reprod Infant Psychol. 2020 Jan 1;38(1):49–59.

46. Rusu PP, Nussbeck FW, Leuchtmann L, Bodenmann G. Stress, dyadic coping, and relationship satisfaction: A longitudinal study disentangling timely stable from yearly fluctuations. PLOS ONE. 2020 Apr 9;15(4):e0231133.

47. Sayal K, Checkley S, Rees M, Jacobs C, Harris T, Papadopoulos A, et al. Effects of social support during weekend leave on cortisol and depression ratings: a pilot study. J Affect Disord. 2002 Sep;71(1–3):153–7.

48. Travis LA, Lyness JM, Shields CG, King DA, Cox C. Social Support, Depression, and Functional Disability in Older Adult Primary-Care Patients. Am J Geriatr Psychiatry. 2004 May;12(3):265–71.

49. Berkman LF. The Role of Social Relations in Health Promotion. Psychosom Med. 1995;57(3):245–54.

50. Ystrom E. Breastfeeding cessation and symptoms of anxiety and depression: a longitudinal cohort study. BMC Pregnancy Childbirth. 2012 Dec 23;12(1):36.

51. Gregory EF, Butz AM, Ghazarian SR, Gross SM, Johnson SB. Are Unmet Breastfeeding Expectations Associated With Maternal Depressive Symptoms? Acad Pediatr. 2015 May;15(3):319–25.

52. Tuthill EL, Pellowski JA, Young SL, Butler LM. Perinatal Depression Among HIV-Infected Women in KwaZulu-Natal South Africa: Prenatal Depression Predicts Lower Rates of Exclusive Breastfeeding. AIDS Behav. 2017 Jun 17;21(6).

53. Avan B, Richter LM, Ramchandani PG, Norris SA, Stein A. Maternal postnatal depression and children’s growth and behaviour during the early years of life: exploring the interaction between physical and mental health. Arch Dis Child. 2010 Sep 1;95(9).

54. Dias CC, Figueiredo B. Breastfeeding and depression: A systematic review of the literature. J Affect Disord. 2015 Jan;171.

55. Tanzania Food and Nutrition Centre. Breastfeeding Support: Close to Mothers: “A Promise Renewed” to promote child survival. Tanzania Food and Nutrition Centre; 2013.

56. De Perre P, Hitimana DG, Simonon A, Dabis F, Msellati P, Karita E, et al. Postnatal transmission of HIV-1 associated with breast abscess. The Lancet. 1992 Jun;339(8807):1490–1.

57. Ndugbu UK, Madukwe CE. Has Knowledge Influenced Right Choice? Assessing the Feeding Practices for Infants of HIV Positive Mothers in a Rural Northern Nigeria. null. 2018;

58. Kapatamoyo B, Andrews B, Bowa K. ASSOCIATION OF HIV WITH BREAST ABSCESS AND ALTERED MICROBIAL SUSCEPTIBILITY PATTERNS.

59. Otoo GE, Lartey AA, Pérez-Escamilla R. Perceived Incentives and Barriers to Exclusive Breastfeeding Among Periurban Ghanaian Women. J Hum Lact. 2009 Feb 1;25(1).

60. Giugliani ERJ. Common problems during lactation and their management. J Pediatr (Rio J). 2004 Nov 1;80(8).

61. Lee S, Kelleher SL. Biological underpinnings of breastfeeding challenges: the role of genetics, diet, and environment on lactation physiology. Am J Physiol Endocrinol Metab. 2016 Aug 1;311(2):E405–22.

62. Young SL, Plenty AHJ, Luwedde FA, Natamba BK, Natureeba P, Achan J, et al. Household Food Insecurity, Maternal Nutritional Status, and Infant Feeding Practices Among HIV-infected Ugandan Women Receiving Combination Antiretroviral Therapy. Matern Child Health J. 2014 Nov 1;18(9):2044–53.

63. Nor B, Ahlberg BM, Doherty T, Zembe Y, Jackson D, Ekström EC. Mother’s perceptions and experiences of infant feeding within a community-based peer counselling intervention in South Africa. Matern Child Nutr. 2012 Oct;8(4):448–58.

64. Laar A. Individual and Community Perspectives, Attitudes, and Practices to Mother-to-Child-Transmission and Infant Feeding among HIV-Positive Mothers in Sub-Saharan Africa: A Systematic Literature Review. Int J MCH AIDS IJMA. 2013;2(1).

65. Ikeako L, Ezegwui H, Nwafor M, Nwogu-Ikojo E, Okeke T. Infant Feeding Practices among HIV-Positive Women in Enugu, Nigeria. Br J Med Med Res. 2015 Jan 10;8(1).

66. Atuyambe LM, Ssegujja E, Ssali S, Tumwine C, Nekesa N, Nannungi A, et al. HIV/AIDS status disclosure increases support, behavioural change and, HIV prevention in the long term: a case for an Urban Clinic, Kampala, Uganda. BMC Health Serv Res. 2014 Dec 21;14(1).

67. Hosseinzadeh H, Hossain SZ, Bazargan-Hejazi S. Perceived stigma and social risk of HIV testing and disclosure among Iranian-Australians living in the Sydney metropolitan area. Sex Health. 2012;9(2).

68. Mebratu L, Mengesha S, Tegene Y, Alano A, Toma A. Exclusive Breast-Feeding Practice and Associated Factors among HIV-Positive Mothers in Governmental Health Facilities, Southern Ethiopia. J Nutr Metab. 2020 Sep 16;2020:1–9.

69. Jones DL, Rodriguez VJ, Mandell LN, Lee TK, Weiss SM, Peltzer K. Influences on Exclusive Breastfeeding Among Rural HIV-Infected South African Women: A Cluster Randomized Control Trial. AIDS Behav. 2018 Sep 20;22(9).

70. Bispo S, Chikhungu L, Rollins N, Siegfried N, Newell ML. Postnatal HIV transmission in breastfed infants of HIV-infected women on ART: a systematic review and meta-analysis. J Int AIDS Soc. 2017;20(1):21251.

71. van Lettow M, Bedell R, Landes M, Gawa L, Gatto S, Mayuni I, et al. Uptake and outcomes of a prevention-of mother-to-child transmission (PMTCT) program in Zomba district, Malawi. BMC Public Health. 2011 Dec 3;11(1).

72. Muyinda H, Seeley J, Pickering H, Barton T. Social aspects of AIDS-related stigma in rural Uganda. Health Place. 1997 Sep;3(3).

73. Futterman D, Shea J, Besser M, Stafford S, Desmond K, Comulada WS, et al. Mamekhaya: a pilot study combining a cognitive-behavioral intervention and mentor mothers with PMTCT services in South Africa. AIDS Care. 2010 Sep 6;22(9):1093–100.

